# Genomic epidemiology of the primary methicillin-resistant *Staphylococcus aureus* clones causing invasive infections in Paraguayan children

**DOI:** 10.1101/2023.01.30.23285178

**Authors:** Fátima Rodríguez, Claudia Salinas, Alejandro Mendoza-Alvarez, Ana Díaz-de Usera, José M Lorenzo-Salazar, Rafaela González-Montelongo, Carlos Flores, Rosa Guillén

## Abstract

2.

Methicillin-resistant *Staphylococcus aureus* (MRSA) is one of the major human pathogens, causing a wide range of infections from food poisoning to necrotizing pneumonia, endocarditis, or septicemia. It could carry numerous resistance genes and virulence factors, some of which are related to the severity of the infection, being also regarded as a potential “Super Bug”. In Paraguay, the prevalence of CC30-ST30-IV clones is the leading cause of *S. aureus* infections both at the regional level and in pediatric population. Here we aimed to deeply analyze the genomic features of MRSA isolates that cause invasive infections in Paraguayan children. An observational, descriptive, cross-sectional study was designed to analyze representative MRSA isolates of the main clones identified between 2009 to 2013 in Paraguayan children. All the genetically linked MRSA isolates were recovered from diverse clinical sources, patients, and hospitals at broad gap periods. Cases were primarily community-acquired, which excludes in-hospital transmission or outbreaks. MRSA isolates were analyzed with short-read paired-end sequencing and assessed for the virulome, resistome, and phylogenetic relationships. The pan-genomic analysis of these clones revealed three major and different clonal complexes (CC8, CC30, and CC5), each composed of clones closely related to each other, despite having different spa types. Furthermore, multiple virulence and resistance genes were identified for the first time in this study, pointing out the complex virulence profiles of MRSA circulating in the country. This study opens a wide range of new possibilities for future projects and trials to improve the existing knowledge on the epidemiology of MRSA circulating in Paraguay.

**Impact statement:** The increasing prevalence of methicillin-resistant *Staphylococcus aureus* (MRSA) is a public health problem worldwide. The most frequent MRSA clones identified in Paraguay in previous studies were (including community and hospital-acquired) the Pediatric (CC5-ST5-IV), the Cordobes-Chilean (CC5-ST5-I), the SouthWest Pacific (CC30-ST30-IV) and the Brazilian (CC8-ST239-III) clones. In this study, the pan-genomic analysis of the most representative MRSA clones circulating in invasive infection in Paraguayan children over ten years (2009-2019), such as the CC30-ST30-IV, CC5-ST5-IV, and CC8-ST8-IV, were carried out to evaluate their genetic diversity, their virulence factors repertoire, and antimicrobial resistance mechanisms revealed multiple virulences and resistance genes pointing out the complex virulence profiles of MRSA circulating in Paraguay. Our work is the first genomic study of MRSA in Paraguay and will contribute to the development of genomic surveillance in the region and our understanding of this pathogen’s global epidemiology.

**Data summary:** The authors confirm all supporting data, code and protocols have been provided within the article or through supplementary data files.

## 5. Introduction

Methicillin-resistant *Staphylococcus aureus* (MRSA), a major human pathogen, has the ability to produce many types of infections, such as food poisoning, necrotizing pneumonia, endocarditis, or septicemia. Initially, MRSA was associated with hospital-acquired infections. However, there has been a considerable increase in cases of healthy individuals who were infected at the community ^1,2^. These infections are particularly problematic due to their associated morbidity, length of hospital stay and mortality ^3^. MRSA could carry numerous virulence factors, some of which explain the severity of the infections it causes, making it a “Super Bug” ^4^.

Nowadays, the complete genome of a microorganism, including the full set of resistance and virulence genes (resistome and virulome, respectively), can be simultaneously characterized by applying next-generation sequencing (NGS) ^5^. In fact, whole genome sequencing (WGS) is instrumental to track the source and propagation of important MRSA clones. Furthermore, WGS is very useful for rapidly infer the emergence of resistance genotypes in the clinical setting to target widely available optimal therapeutic approaches, especially during invasive infection treatments ^6^.

Severe infections caused by methicillin-resistant strains acquired from the community (CA-MRSA) in the United States at the beginning of the century, called USA300, were reported, which quickly dispersed geographically, displacing less virulent strains, and becoming the leading cause of skin and soft tissue infections (SSTI) in this country ^7–9^. The most frequent MRSA clones (including community and hospital-acquired) in South America are the Pediatric (CC5-ST5-IV), the Cordobes-Chilean (CC5-ST5-I), the SouthWest Pacific (CC30-ST30-IV), the Brazilian (CC8-ST239-III), and New York/Japan (CC5-ST5-II) clones ^10^. In Argentina, the main CA-MRSA clone related to invasive infections in the last decade is CC30-ST30-IV-t019 PVL+, which became predominant, replacing the previously described CC5-ST5-IV-t311 PVL+ ^11–13^. In Paraguay, the prevalence of CA-MRSA CC30-ST30-IV clone is the leading cause of *S. aureus* infections both at the regional level and in pediatric population ^11,14,15^. The objective of this study was the deep characterization of the genomic features of MRSA isolates causing invasive infections in Paraguayan children.

## 6. Methods

### Bacterial strains

An observational, descriptive and, cross-sectional study was designed to analyze representative MRSA isolates of the main clones identified between 2009 to 2013, from the Microbiology Department of the Health Sciences Research Institute, UNA, Paraguay Biobank (maintained at -80 ºC in BHI+20% glycerol). Isolates were initially recovered from invasive infections of children under sixteen years old attending any of the four reference hospitals from Asunción and the Central Department of Paraguay, from different clinical specimens. Identification data, epidemiology files, and records of antimicrobial susceptibility were extracted from the epidemiological record of the isolates. Phenotypic identification of the isolates and the antimicrobial susceptibility tests were carried out following the criteria recommended by the CLSI (Clinical and Laboratory Standards Institute) from 2009 to 2013, according to the strain collection date or by automated systems using Vitek^®^2 (BioMérieux, La Balme, French) according to the manufacturer’s instructions. These strains were sub-cultured in Tryptic Soy Agar (TSA, Difco, Le Pont de Claix, France) medium from primary cultures and incubated for 24 h at 35 °C under 5% CO_2_ for further molecular characterization.

### DNA extraction and genotyping

Total bacterial DNA was extracted from pure MRSA cultures using the Wizard^®^ Genomic DNA Purification kit (Wizard Genomic, Promega, Madison, USA) following the manufacturer’s instructions. MRSA was molecularly typed by spa typing ^16^ and multi-locus sequence typing (MLST) ^17^. Detection of *mecA* and Panton-Valentine leukocidin (PVL) coding genes were carried out as described previously ^18^, and the characterization by multi-locus variable analysis (MLVA) was carried out by a multiplex PCR as described elsewhere ^19^. The *staphylococcal cassette chromosome mec* (*SCCmec*) element was typed using the Kondo’s typing system ^20^.

### Library preparation and whole-genome sequencing

All samples were purified and concentrated previously to library preparation with DNA Clean & Concentrator (Zymo Research), following the manufacturer’s recommendations. Dual index libraries were processed with Nextera XT DNA Library Preparation Kit (Illumina Inc., California, USA) following the manufacturer’s recommendations with manual library normalization. Paired-end sequencing was performed on a MiSeq Sequencing System (Illumina Inc., California, USA) with 300 bp reads to a theoretical sequencing throughput of 3 Mb/library (minimal expected coverage of 100X). The library concentration was loaded at 10 pM, and 5% of PhiX Control V3 was used as the internal control. Sequencing was conducted in the Genomic Division of the Instituto Tecnológico y de Energías Renovables (ITER, Tenerife, Spain).

### Bioinformatic analysis

BCL files were converted to demultiplexed FASTQ files using BCLtoFASTQ v2.19 tool. Quality control was performed with FastQC v0.73 ^21^ to assess sequencing qualities, read length, and the total number of reads. Taxonomic correspondence (species identification) was obtained with Kraken v2.1.1 ^22^. Then, the reads were subjected to a trimming process to improve their quality using the Trimmomatic software v0.38.1 ^23^. Subsequently, de novo assembly was carried out with Unicycler v0.4.8.0 ^24^ and its quality control with Quast Genome assembly Quality v5.0.2 ^25,26^ to assess the quality of the assembly. A summary report was obtained with assembly metrics such as total genome size, total number of contigs, largest contig size, and contig with size greater than 1Kb, N50, and GC content. All assemblies are available at the NCBI Sequence Read Archive (BioProject accession number PRJNA830493). Finally, the assemblies were processed with Prokka (Prokaryotic Genome Annotation) v1.14.6 for bacterial annotation ^27^. All the analyses were conducted on the TeideHPC Supercomputing facility (http://teidehpc.iter.es).

Additional characterization of the isolates was carried out using the assemblies in combination with the software MLST (PubMLST database) v2.19.0 ^28,29^ for detecting the sequence type, and the Center for Genomic Epidemiology (CGE) platform with spaTyper software v1.0 for spa type identification ^30^. ABRicate v1.0.1 was combined with different databases, such as ResFinder and Bacterial Antimicrobial Resistance Reference Gene NCBI, to detect antibiotic resistance genes, the possible induction of resistance, and the detection of virulence factors genes with the Virulence Factor Data Base (VFDB) ^31^. For detecting the genes encoding virulence factors, we also analyzed the genomic assemblies in the CGE platform in combination with the Virulence finder software v2.0.3. We use the NG-CHM Builder: Cluster Matrix platform from the University of Texas (https://build.ngchm.net/NGCHM-web-builder/) to analyze the virulence factors profiles detected.

Finally, a phylogenetic analysis was performed from the assembled bacterial genomes using Roary v3.13.0, IQ-Tree v1.5.5, and FigTree v.1.4.4 (http://tree.bio.ed.ac.uk/software/figtree/), in order to establish the core genome of the clinical isolates analyzed ^32,33^. The pan-genomic analysis was carried out using only the SNP alignment with the SNP sites v2.5.1, and a Newick phylogenetic tree was derived to depict the relationships between each strain ^34^. The output files generated by Roary software were visualized in the online tool Microreact https://docs.microreact.org/ ^35^, generating the Newick-based tree with the genotypes and demographic characteristics of the isolates. The reference strains used in the pan-genomic analysis were as follows: MRSA 252 (NC_002952.2), TCH60 (NC_017342.1), NCTC8325 (NC_007795.1), N315 (NC_002745.2), and USA300 (NZ_CP092052.1).

All the bioinformatics software used for this study was run using default parameters.

## 7. Results

The Microbiology Department biobank currently has 952 isolates of *S. aureus*, of which 262 are MRSA, and 84 of them have caused invasive infections. The ten isolates analyzed in this study represent the majority of the 84 MRSA invasive profiles. Clinical and genotypic characteristics are shown in **table 1**. All MRSA isolates (100%, 10/10) came from patients, both male and female, under 15 years of age. Regarding susceptibility to antibiotics, all MRSA isolates (100%, 10-10) were resistant to penicillin and sensitive to vancomycin, gentamicin, ciprofloxacin, trimethoprim-sulfamethoxazole, and have the *mecA* gene and the *staphylococcal cassette SCCmec* IV. The sequence type and spa type detected by WGS were fully concordant with the results obtained by PCR **(Table 1)**. This study includes two isolates characterized as CC8-ST8-IV, which caused sepsis in children in two healthcare centers in Paraguay between 2012 and 2013.

**Table 1.** Clinical, phenotypic, and molecular features of MRSA selected for next-generation sequencing (n=10).

| Sample ID | Collection date | Sample | Patient disease | CC | MLST | Spa type | MLVA | *lukF/S-PV* |
| --- | --- | --- | --- | --- | --- | --- | --- | --- |
| **SGP_11** | 2009, Dec | Blood | Pneumonia | 30 | 30 | t019 | 1 | + |
| **SIP_29** | 2010, Jan | Blood | Osteomyelitis | 30 | 30 | t019 | 1 | + |
| **SGP_29** | 2010, Feb | Blood | Sepsis | 5 | 5 | t311 | 2 | + |
| **SGP_63** | 2011, Nov | Blood | Sepsis | 8 | 8 | t11770 | 4 | + |
| **SGP_102** | 2012, Jan | Blood | Sepsis | 30 | 30 | t019 | 1 | + |
| **SCM_71** | 2012, Feb | Purulent discharge | Sepsis | 5 | 100 | t002 | 3 | - |
| **SCM_77** | 2012, Apr | Purulent discharge | Sepsis | 5 | 5 | t311 | 2 | + |
| **SHN_80** | 2012, Nov | Blood | Sepsis | 30 | 30 | t021 | 1 | - |
| **GIP_4** | 2013, Aug | Blood | Osteomyelitis | 8 | 8 | t400 | 5 | + |
| **GIP_64** | 2013, Dec | Blood | Pneumonia | 30 | 30 | t019 | 1 | + |

**Abbreviations:** ID: Identification, CC: Clonal complex, MLST: Multi-locus sequence type, MLVA: Multi-locus variable analysis, *lukF/S-PV*: Panton Valentine leukocidin gene, Jan: January, Feb: February, Apr: April, Aug: August, Nov: November, Dec: December.

According to WGS, all the isolates analyzed carried the *mecA* resistant gene, which confers resistance to methicillin. The susceptibility profile and resistome to other antibiotics is shown in **table 2**. For all resistance genes, the sequence identity percentages ranged between 97.1-100.0% and had a breath of coverage between 87.2-100.0%. The eighty-seven virulence factor genes detected are shown in **figure 1**. The toxin and virulence gene content were diverse and correlated with the typing characteristics **(Figure 1)**. All isolates carried aureolysin (*aur*), gamma-hemolysin (*hlg*) A, B, and C components, iron-regulated surface determinant protein (*isd*) A, B, C, E, F, and G components, alpha-hemolysin (*hly/hla*), staphylokinase precursor (*sak*), cell surface elastin (*ebp*), and clumping factor A fibrinogen binding protein (*clfA*). In addition, all of them carry the IgG and IgA binding protein *(spa)*, complement inhibitor SCIN (*scn*), glycerol ester hydrolase *(geh)*, the protein secretion system components *(es)* EsxA, EsaA/B, and EssA/B and, the type 8 capsular polysaccharide synthesis components *(cap8)* L/M/N/O/P, with antiphagocytic functions. The serine protease *(sspA)*, staphopain, cysteine proteinase *(sspB)*, staphostatin B *(sspC)*, and NPQTN specific sortase B *(srtB)* were also detected in all isolates. The differential presence of the other virulence genes detected by WGS is shown in **table 3** for a better understanding and classification according to their functionality. None of the sequenced isolates carried enterotoxins (*se*) B, C, D, H, L, and exfoliative toxins (*et*) A/B. Whole genome phylogenetic analysis clustered the isolates into three distinct clades: CC30-ST30 (SGP_11, SIP_29, SGP_102, GIP_64 and SHN_80), CC5-ST5/ST100 (SGP_29, SCM_71 and SCM_77), and CC8-ST8 (SGP_63 and GIP_4). Its geographical and timeline distribution is showed in **figure 2**.

**Table 2.**
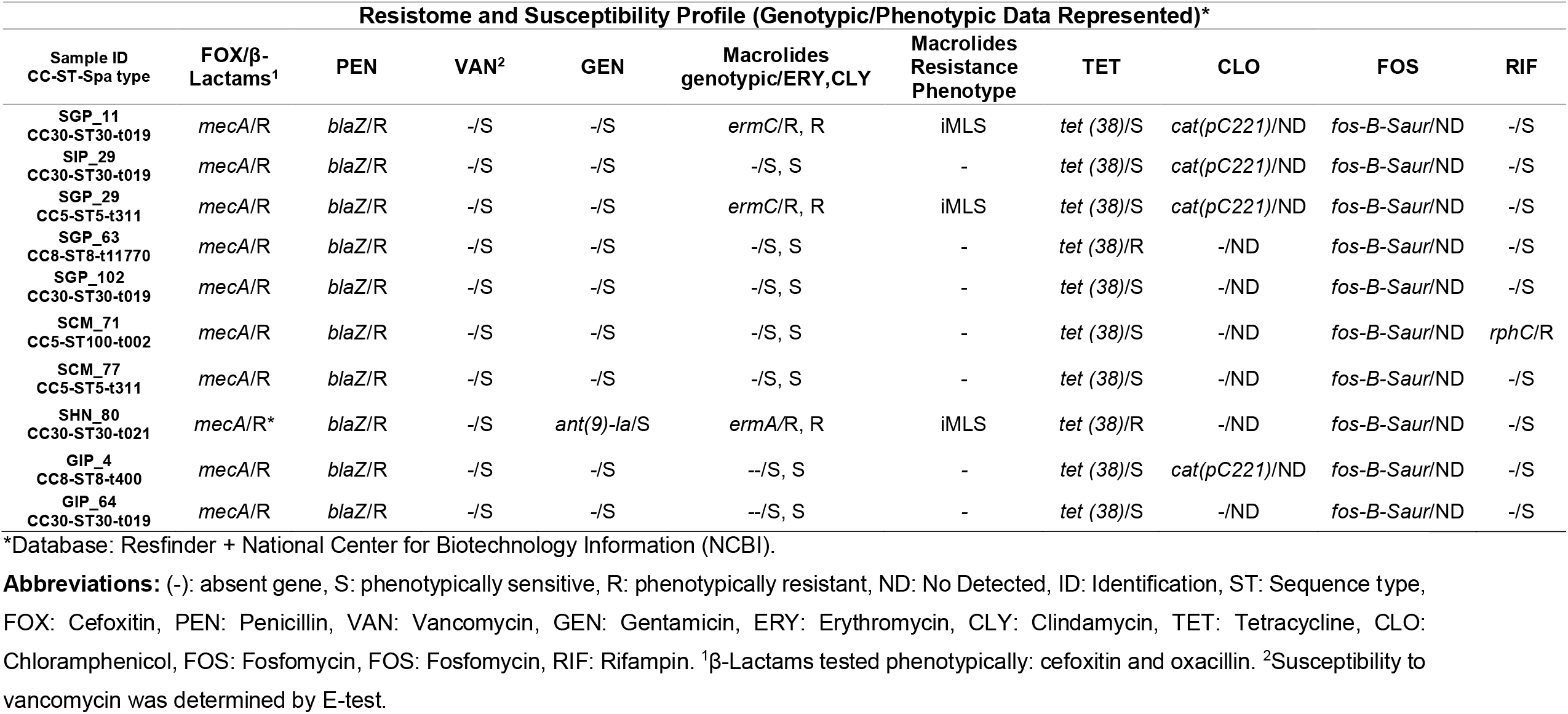
Resistance genotypes and phenotypes identified in the MRSA isolates through the genome analysis (n=10).

**Table 3.**
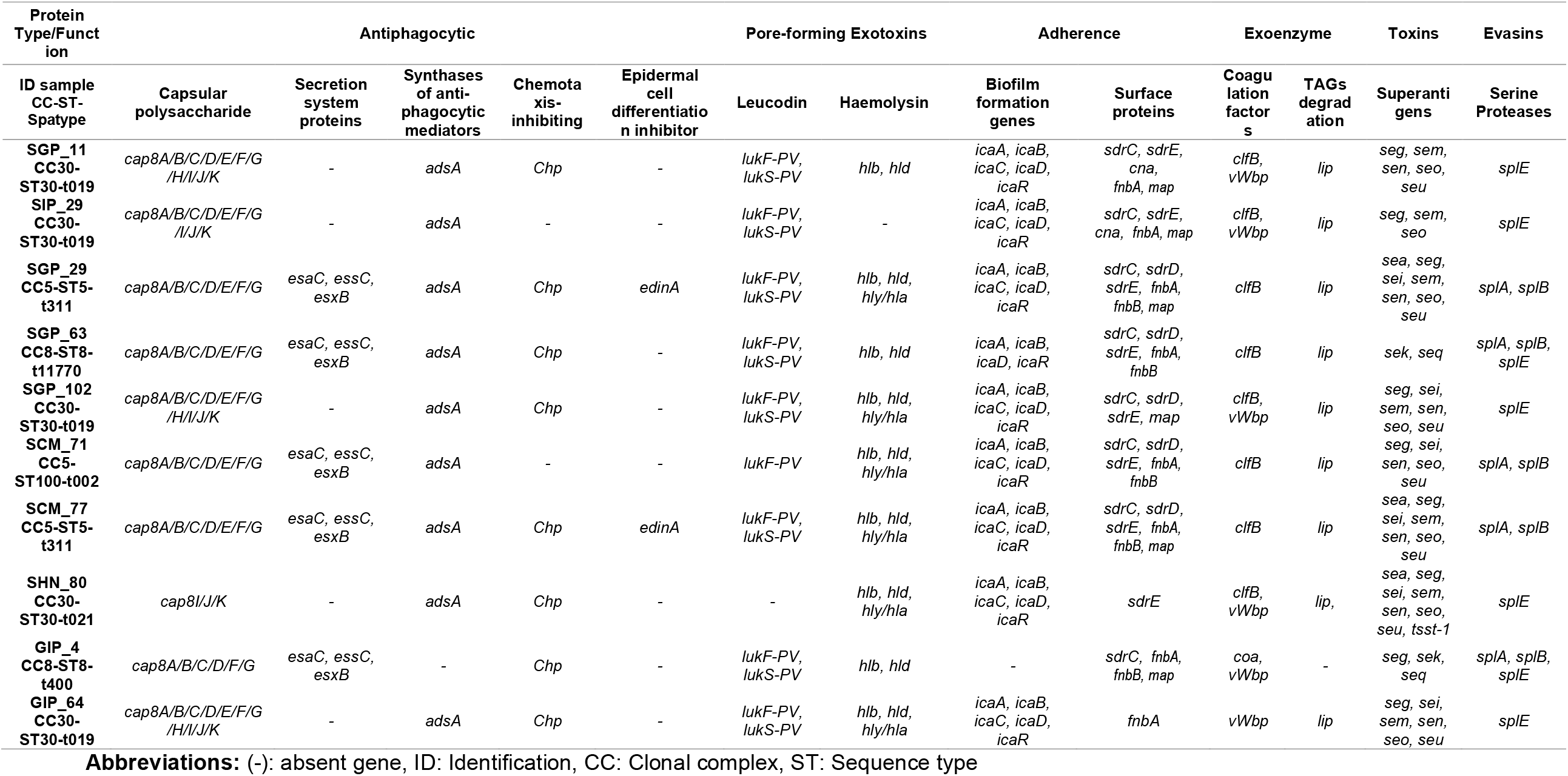
Virulome of the sequenced MRSA isolates (n=10).

**Figure 1.**
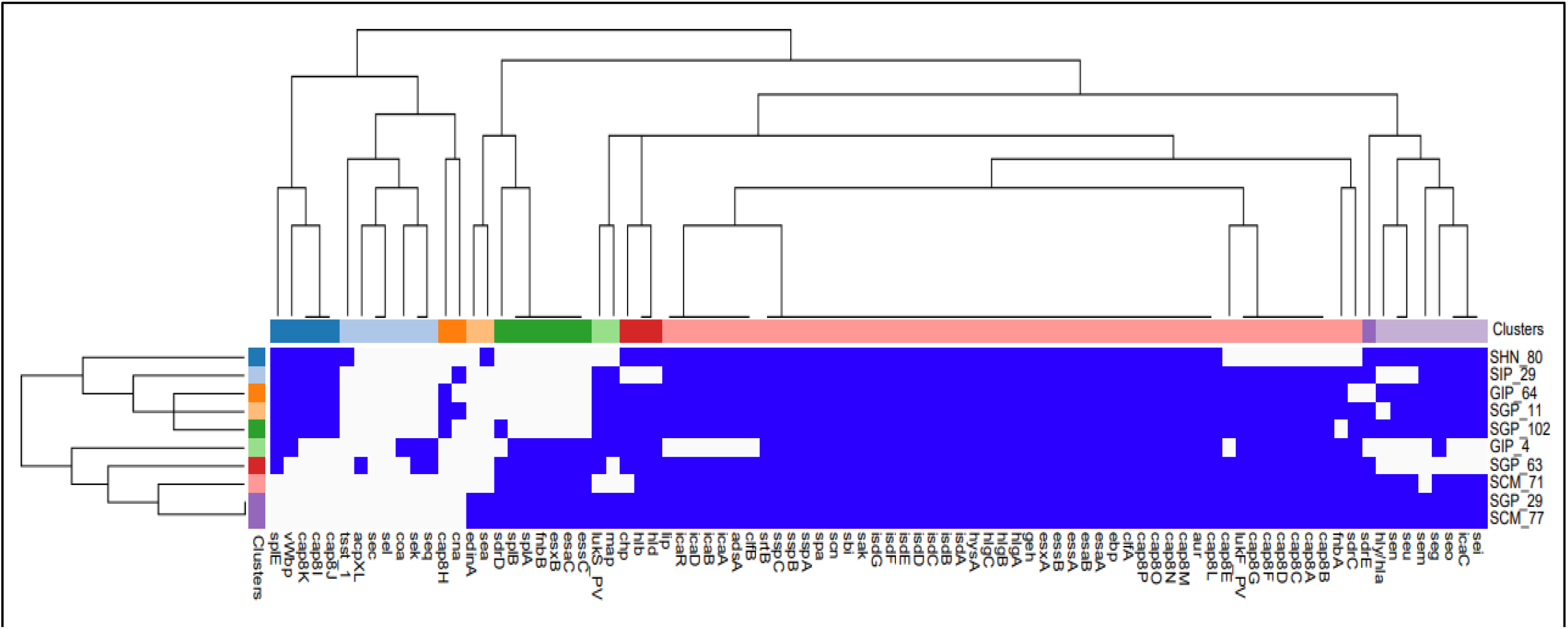
Virulence factors Heat map and dendrogram from the de novo assembly of MRSA genomes (n=10). Blue boxes indicate the presence of each one of the virulence factor encoding genes analyzed (n=87). The isolates were clustered hierarchically based on their virulence factor profile using Euclidean metric distance with complete linkage clustering in both rows and columns, thus providing two dendrograms. The top dendrogram (top) clusters virulence factor clustered according to their frequency in the isolates: the most frequent: in the middle and the least frequent in the extremes. The left dendrogram (left) clustered the isolates in terms of their similarity in virulence profile; within the same cluster are **SIP_29, GIP_64, SGP_11**, and **SGP_102**, all **CC30-ST30-t019-IV** and **SHN_80** a little further away **CC30-ST30-t021-IV**. In another cluster as identical to **SCM_77** and **SGP_29** (both **CC5-ST5-t311-IV**), then to **SCM_71 (CC5-ST100-t002-IV), SGP_63 (CC8-ST8-t11770-IV)** and **GIP_4 (CC8-ST8-t400-IV)**.

**Figure 2:**
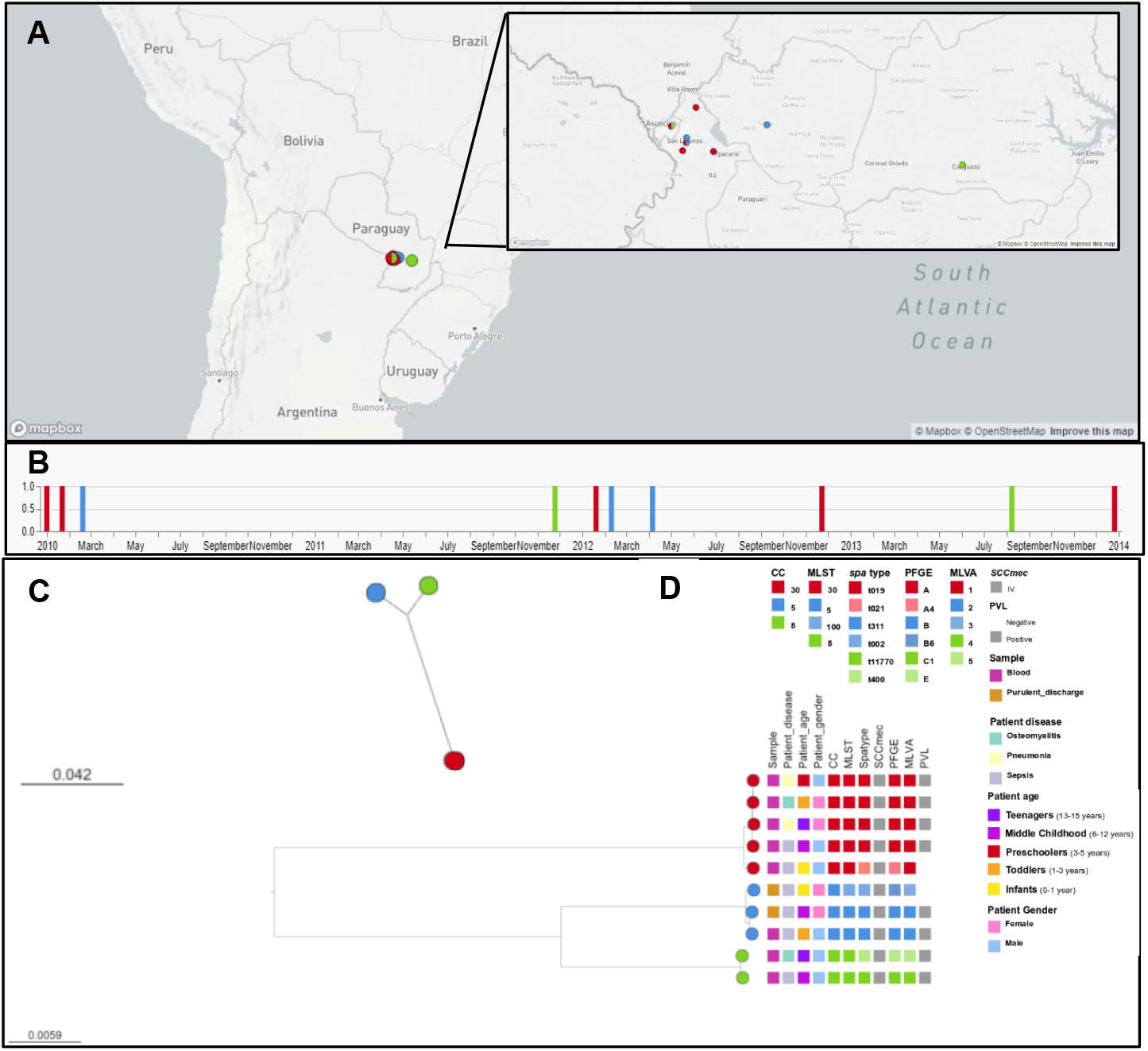
Phylogeny, molecular and demographic characteristics of MRSA that cause invasive infection in Paraguayan children (n=10). (A) Geographical distribution of samples taken in Paraguay; (B) Timeline of samples used in this study; (C) Midpoint-rooted phylogenetic tree inferred from 35,402 SNP sites obtained after aligning the ten genomes in study (CC30, CC5 and CC8). Tree branches and nodes are colored by clonal complex (CC) of strain as well as indicated on (A) and in (B). The distribution of genotypes and demographic characteristics are shown as tree metadata blocks (D). Data are available at: https://microreact.org/project/3ws3rcf2vdzkzhcvpnxmx8.

## 8. Discussion

Despite the burden of MRSA infection in Paraguay, this constitutes the first WGS analysis on MRSA isolates to be completed in the region. For that, we included the most representative MRSA clones circulating in invasive infection in Paraguayan children, such as the CC30-ST30, CC5-ST5, and CC8-ST8 ^14^, to evaluate their genetic diversity, their virulence factors repertoire, and antimicrobial resistance mechanisms.

MRSA clones represent a significant health concern worldwide. There is a clear need to understand better their genomic structure, transmission dynamics, and evolution in a geographic region over time ^1^. The MRSA post-genomic era revealed new virulence factors and antimicrobial resistance mechanisms ^6^. The MRSA clones identified as the main circulating in Paraguay in previous studies by our research group were CC30-ST30-t019-IV (77%) and CC5-ST5-t311-IV (10%) in MRSA isolates collected between the years 2009 and 2013 in diverse infectious processes (SSTIs and invasive) in Paraguayan children ^14^. In the period 2010-2011, the CC5-ST5-IV lineage (54%) was firmly installed in the community. Over the years, it was widely displaced by the CC30-ST30-IV. The clonal complex 30 (30) groups pandemic *S. aureus* lineages have been consistently reported from all continents, including CC30-ST30-IV, one of the most prevalent MRSA lineages in Argentina and Paraguay ^11,14,36^. This well-known virulent strain caused skin lesions, sepsis, and pneumonia in children and young adults in hospitals and the community. In recent descriptive studies, this clone has been associated with an increased risk of infective endocarditis ^37^ and persistent bacteremia due to *S. aureus* ^38^.

In the present study, all the MRSA analyzed carried the *cassette SCCmec IV*, which is smaller than the others and has a simpler genetic makeup, carrying only methicillin resistance genes (*mecA*) ^1^. Regarding the resistome analysis, most MRSA showed phenotypic and genotypic concordance in antibiotic susceptibility results (89%). Similar findings were reported in other studies that showed concordance in 76-100% of isolates ^39,40^. Our results showed discordance between genotype and phenotype in isolates that carried genes resistant to tetracycline (80%) and gentamycin (10%). This could be due to mutations in these resistance genes or defects in their expression that should be further explored experimentally in the future.

We also obtained data for other antimicrobials that are not routinely tested in the laboratory, such as fosfomycin and chloramphenicol, detecting their potential resistance in 100% (10/10) and 40% (4/10) of MRSA isolates, respectively. Fosfomycin is not used to treat staphylococcal infections in Paraguay and Argentina. This is the reason why it was not included in the antibiotic panel. Surprisingly, all strains were predicted to be resistant to fosfomycin, carrying the acquired f*ofB* gene ^41^. Fosfomycin is a broad-spectrum agent frequently active against antimicrobial-resistant bacterial pathogens, including MRSA. It is a bactericidal agent that inhibits cell wall synthesis. It is most frequently used with other antimicrobial agents (e.g., *β*-lactams, carbapenems, and aminoglycosides) and has an excellent safety profile, including in neonates and children, even with long-term effects administration (i.e., weeks). Fosfomycin has the ability to reach a wide range of tissues and it could be used for the most common infections, such as bacteremia, urinary tract, skin and soft tissue, and respiratory infections, and for infections difficult to treat, such as bone meningitis and invasive ocular infections. It has been suggested that fosfomycin could be reused to treat MRSA MDR (multi-drug resistant) infections that have not responded to first- and, potentially, second-line antimicrobials ^42^. Chloramphenicol is another broad-spectrum antibiotic that had been partially abandoned in developed countries because since its systemic administration is associated with fatal aplastic anemia but it is now widely used in the world due to the continuing problem of MDR pathogens ^43^. Our findings, although only based on WGS data without experimental validation, indicate that fosfomycin and chloramphenicol would not be optimal therapeutic options for treating MRSA infections in Paraguay and probably in the region because of the potential resistance presented by the main MRSA clones that circulate in the country.

Regarding the virulome of the isolates, we identified the co-existence of multiple virulence factor groups. The virulence factors detected in all of the MRSA analyzed were pore-forming exotoxins (aureolysin, alpha, and gamma-hemolysin A, B, and C components) ^44^, potent plasminogen activator (staphylokinase), cell wall-anchored surface proteins (protein A, serine protease operon, cell surface elastin, clumping factor A and, iron-regulated surface determinant protein A, B, C, E, F, and G components) ^45–47^. We also detected genes encoding proteins with antiphagocytic functions (type 8 capsular polysaccharide, IgG and IgA binding, complement inhibitor SCIN, glycerol ester hydrolase, and part of the secretion system components EsxA, EsaA/B, and EssA/B), spreading factor protein (hyaluronidase) ^48^, immunoglobulin binding protein (*sbi*) ^49^, and a family of membrane-associated bacterial enzymes (sortases) ^50^. They all have a vital function for survival, reproduction, colonization, and bacterial spread, which could explain why they are part of the *S. aureus* core genome.

The main MRSA clones circulating in Paraguay show differences in the virulence profiles. CC30-ST30-t019-IV is characterized by having the largest number and diversity of genes encoding capsular polysaccharides, which expresses proteins with antiphagocytic activity and inhibits the interaction between C3b, immunoglobulin, and receptors. This clonal complex (CC30) has gained genes that encode the coagulation factor protein (*vWbp*) and superantigens (SAgs) encoded in the EGC cluster (G, I, M, N, O, U), and the toxic shock syndrome toxin (*tsst-1*), all of which are involved in aggravating the infectious process ^51–53^. The superantigens (SAgs) encoded in the EGC cluster (G, I, M, N, O, U) are currently considered the most prevalent staphylococcal toxins among clinical and colonizing isolates. They are present in 50-70% of nasal carriers and its expression may be crucial for the development and aggravation of some infections, such as respiratory and endocarditis ^51,54^. Experimentally, it has been shown in animal models that the expression of the EGC cluster and *tsst-1* genes together contributes to higher mortality and/or a more rapid and complicated progression of the infection, generally accompanied by lethal complications such as heart failure and cerebrovascular accidents. Even so, clinical infections progress in the presence of EGC cluster toxins, even without the systemic or local effects of TSST-1. Therefore, it is postulated that the EGC cluster toxins are also responsible for the worsening of the infectious, probably on a smaller scale than TSST-1 ^55^.

CC30-ST30-IV and CC8-ST8-IV clones win the serine proteases *splE* genes, encoding evasins with a possible cooperative and complementary activities with the other *spl* proteases (lost by CC30), apparently constitute an extracellular digestive system with a role in the pathogenesis of *S. aureus* ^55^. In contrast, the loss of the secretion system proteins used by bacteria to interact and manipulate their environments is significant for adhesion and permanence in the host-cell ^56^, as well as the epidermal cell differentiation inhibitor (*edinA*), which is involved in the bacterial dissemination process and hindering complement-mediated phagocytosis ^57^. This lack of genes could give these lineages (CC30 and CC8) better efficiency for propagation due to their smaller size and the lower fitness costs associated with carrying fewer genes, choosing those involved in the evasion of the immune response, infection expansion, and propagation process, and being an explanation of it displacing the other lineages ^1^.

The presence of a new clone of the CC30-ST30-IV lineage, related to the most frequently reported CC30-ST30-IV-t019, but with spa-type (t021), shows that this clone begins to differentiate in the *spA* gene sequences, in this case in three repetitions by insertion and deletion. This phenomenon generally appears in chronic or repeated infections, indicating that clone CC30-ST30-t019 is probably beginning to change, evolving to other spa-types ^15^.

The CC5-ST5-t311-IV clones study here (SGP_29 and SCM_77), carried the enterotoxin A (*seA*), PVL (*lukF/S-PV*), and superantigens (SAgs) encoded in the EGC cluster (G, I, M, N, O, U) like a signature of highly toxic strains, involved in CA-MRSA infections. This is even more significant because these isolates come from invasive infections. The SCM_71 strain, CC5-ST100-t002-IV, and resistant to rifampicin, known as the pediatric clone, is highly related to CC5-ST5-t311-IV and differs from it by a point mutation in the *aroE* gene (substitution of the aroE4 allele, characteristic of ST5, by the aroE65 characteristic of ST100), as well as by the insertion of a repeat r17 in the fifth position of the *spA* gene that differentiates it from t311 and converts it into spa-type t002 (58). The CC5-ST100-t002-IV clone could be a pathway on the CC5-ST5-t311-IV evolution, allowing its adaptation to the sanitary conditions of the region, such as antibiotic pressure due to the use of rifampicin in the treatment of SSTIs in the community, which is sustained by the resistance to rifampicin that this clone presents ^58^.

We identified the potential ability to biofilm formation in almost all the MRSA isolates analyzed, except in the CC8-ST8-t400-IV clone. The biofilm is another crucial factor contributing to staphylococcal infections, implicated in various persistent human microbial infectious diseases, allowing them to evade multiple clearance mechanisms, such as antimicrobials and the host immune system leading to treatment failure and recurrent/chronic infections. MRSA biofilm and virulence factors production are closely linked since the primary biofilm regulator, the accessory gene regulator *agr*, is also vital for expressing numerous virulence factors. Therefore, many biofilm-related virulence factors have been the target of research on *S. aureus* therapeutics ^50^. Tree of the ten MRSA isolates studied here can form biofilms with moderate intensity, showed by in vitro testing assays. Some of them could be inhibited in the presence of two methanolic extracts from Paraguayan native plants called *Pterocaulon alopecuroides* and *Pterocaulon angustifolium* ^59^. The detection of biofilm formation, especially in chronic infections, is crucial since it affects the choice of strategies for their elimination, such as surgical removal, given the ineffectiveness of traditional antibiotic therapies ^59,60^.

The CC8-ST8-lV clones (t11770 and t400) carried the serine proteases system complex (splA/B/E), a digestive system extracellular with a role in the pathogenesis of MRSA, PVL, and the enterotoxins K, Q, characteristics from CC8 complex like a superbug, among others, all of which contributing to their virulence profile ^1^. These strains are closely related to the CC8-ST8-IV-PVL+ clone, identified as the major cause of CA-MRSA infections in North America, which have lately been increasingly reported worldwide ^8^.

All the genetically linked MRSA isolates were recovered from diverse clinical sources, patients, and hospitals at broad gap periods. They were primarily community-acquired, which excludes in-hospital transmission or outbreaks. The pangenomic analysis of these clones revealed three major and different clonal complexes (CC8, CC30, CC5), each composed of clones closely related to each other, despite having different spa types.

In conclusion, the use of WGS in the present study added value to the classic isolate identification and molecular typing protocols. It offered precious and precise genomic data about the most prevalent MRSA clones circulating in the country. Multiple virulence and resistance genes were identified for the first time in this study in samples from the country, indicative of the complex virulence profiles of MRSA that are circulating in Paraguay. This critical qualitative leap opened a wide range of new possibilities for future projects and trials to improve the existing knowledge on the epidemiology of MRSA circulating in Paraguay.

## Data Availability

All assemblies are available at the NCBI Sequence Read Archive (BioProject accession number PRJNA830493).

https://www.ncbi.nlm.nih.gov/bioproject/?term=PRJNA830493

## 9. Author statements

### 9.1 Author contributions

F.R. conceptualization, data curation, formal analysis, investigation, resources, software, validation, visualization, writing-original draft and writing-review and editing. C.S. investigation and writing-review and editing. A.M.A. formal analysis, software, and writing-review and editing. A.D.U. formal analysis and writing-review and editing. J.M.L.S. resources and writing-review and editing. R.G.M. investigation, validation and writing-review and editing. C.F. resources and writing-review and editing. R.G. conceptualization, funding acquisition, investigation, project administration, resources, software, visualization writing-review and editing.

### 9.2 Conflicts of interest

The authors declare that there are no conflicts of interest.

### 9.3 Funding information

The present study was carried out through an R&D stay co-financed by CONACYT of Paraguay through the Program for Linking Scientists and Technologists, call 2018 (PVCT18-61) with FEEI resources.

AM-A was supported by a fellowship from the Canarian Agency for Research, Innovation and Information Society (ACIISI, Grant No. TESIS2020010002) co-funded by the European Social Fund (ESF).

### 9.4 Ethical approval

This study was approved by the Ethics and Scientific Committee of the Institute of Research in Health Sciences (P20/2011, P44/2012). The samples were anonymized and analyzed according to the local regulations and laws that apply to medical information.

## 9.5 Acknowledgements

The authors acknowledge the support of the Freiburg Galaxy Server (Germany) funded by the Collaborative Research Centre 992 Medical Epigenetics (DFG grant SFB 992/1 2012) and the German Federal Ministry of Education and Research BMBF grant 031 A538A de.NBI-RBC.

## Notes

### Competing Interest Statement

The authors have declared no competing interest.

### Author Declarations

This study was approved by the Ethics and Scientific Committee of the Institute of Research in Health Sciences of Paraguay (P20/2011, P44/2012).

